# Escape of SARS-CoV-2 variant Omicron to mucosal immunity in vaccinated subjects

**DOI:** 10.1101/2022.05.03.22274517

**Authors:** Emanuela Martinuzzi, Jacques Boutros, Jonathan Benzaquen, Nicolas Glaichenhaus, Paul Hofman, Charles Hugo Marquette

## Abstract

Omicron s escape to vaccine-induced systemic antibody responses has been shown in several studies in Omicron-infected patients and vaccine controls.

In the present study we compared mucosal antibody response to Omicron to mucosal antibody response to ancestral strain and Delta variant. This was done on nasal epithelial lining fluid (NELF) prospectively collected in 84 otherwise healthy healthcare workers who had never exhibited PCR-documented COVID-19 and had received three doses of the Pfizer-BioNTech COVID-19 mRNA vaccine. NELF was collected prior to Omicron detection in the geographical area of inclusion.

We show that NELF antibodies from vaccinated individuals were less efficient at inhibiting the binding of the Omicron Spike protein to ACE-2 compared to those of Delta or the ancestral strain.

These findings may explain the increased risk of onward transmission of Omicron, consistent with its successful global displacement of Delta in countries with a high vaccination coverage.

---

Since it was first reported on November 24^th^ 2021 and classified as a variant of concern two days later ^1^, the Severe Acute Respiratory Syndrome Coronavirus 2 (SARS-CoV-2) variant Omicron (B.1.1.529) has spread throughout the world ^1,2^, rapidly replacing other circulating variants including Delta. The growth advantage of Omicron over Delta was observed not only in countries with low vaccination coverage such as South Africa, but also in those in which more than 90% of individuals where vaccinated such as Norway ^3^. In keeping with the higher transmissibility of Omicron, two recent studies in English and Norwegian households have shown that the secondary attack rate was higher when the variant of the index case was Omicron compared to Delta ^4,5^.

Compared to the ancestral reference strain, Omicron harbors 60 mutations among which 32 in the gene of the Spike protein. Some of these mutations result in a higher affinity of Spike for its human angiotensin-converting enzyme 2 (ACE2) receptor, a phenomenon that explained at least in part why Omicron multiplies in the upper airways 70 times faster than Delta ^6^. Mutations in Spike also account for the ability of Omicron to escape systemic antibody responses as demonstrated by the reduced ability of plasma antibodies from both COVID-19 convalescent or vaccinated subjects to bind the Omicron Spike protein and to inhibit its binding to ACE-2 ^7,8^.

While protection from severe forms of COVID-19 in vaccinated adults is mediated at least in part by SARS-CoV-2-specific antibodies, lessons drawn from other mucosal pathogens suggest that mucosal antibodies and especially secretory immunoglobulin A (sIgA) are those that efficiently block transmission of respiratory viruses such as SARS-CoV-2 ^9^. Therefore, we hypothesize that Omicron disseminates more rapidly than Delta in vaccinated subjects because it escapes vaccine-induced mucosal immune responses.

To test this hypothesis, we prospectively collected nasal epithelial lining fluid (NELF) in a cohort of 84 otherwise healthy healthcare workers (Centre Hospitalier Universitaire de Nice) who had never exhibited PCR-documented COVID-19 and had received three doses of the Pfizer-BioNTech COVID-19 mRNA vaccine 10 to 131 days before NELF collection. In all subjects NELF was collected prior to Omicron detection in the geographical area of inclusion, i.e. between December 14 and December 31 2021. All subjects signed an informed consent to participate in this work. This study was approved by the CPP Sud Méditerranée V ethics committee (ClinicalTrial.gov identifier: NCT04418206).

NELF was gently collected with Hydroxylated polyvinyl acetate (PVA) sponges (Merocel^®^ Standard Dressing, ref 400400, Medtronic, Minneapolis, MN, US), inserted between the nasal septum and the inferior turbinate, left in place for 15 minutes until they swelled, gently retrieved and placed in a 50 ml Falcon tube (Dustcher, Bernolsheim, France) containing 2 ml of saline solution. The fluid contained in the sponge (saline + nasal secretions) was then extracted by simple pressure, aliquoted and frozen at -70°C until further analysis. Serum albumin concentration in NELF was more than 100-fold lower in NELF compared to serum ruling out the possibility that collecting nasal fluids using swabs could have irritated the epithelia eventually increasing exudation of tissue fluids. IgA and IgG to Spike of the SARS-CoV-2 ancestral strain and of its Delta and Omicron variant were measured using the V-PLEX® SARS-CoV-2 Panel (MSD, Maryland, US), as described ^10^. Total IgA and IgG levels were measured using the V-PLEX® Isotyping Panel 1 Human/NHP Kit. Nasal fluids were diluted 10-fold before being assessed for Spike-specific and total IgA and IgG. Data were acquired on the V-PLEX® Sector Imager 2400 plate reader and analyzed using Discovery Workbench 3·0 software. NELF antibodies were assessed for their ability to inhibit the binding of a soluble ACE2 to Spike of the ancestral SARS-CoV-2 strain and its Delta and Omicron variants using the multiplex V-PLEX® SARS-CoV-2 Panel 13 ACE2 Kit as described ^10^. NELF were diluted 10-fold before being assessed for binding inhibition. Data were acquired on the V-PLEX® Sector Imager 2400 plate reader and analyzed using the Discovery Workbench 3·0 software (MSD). Diluent alone was used as a blank. The percentage inhibition was calculated according to manufacturer’s instructions. This assay has been shown to correlate with assays for viral neutralization.

We found that mucosal IgG and IgA bound less efficiently to the Omicron Spike protein compared to those of Delta or the Wuhan ancestral strain (Figure 1A, 1B). NELF antibodies from vaccinated individuals were also less efficient at inhibiting the binding of the Omicron Spike protein to ACE-2 compared to those of Delta or the ancestral strain (Figure 1C).

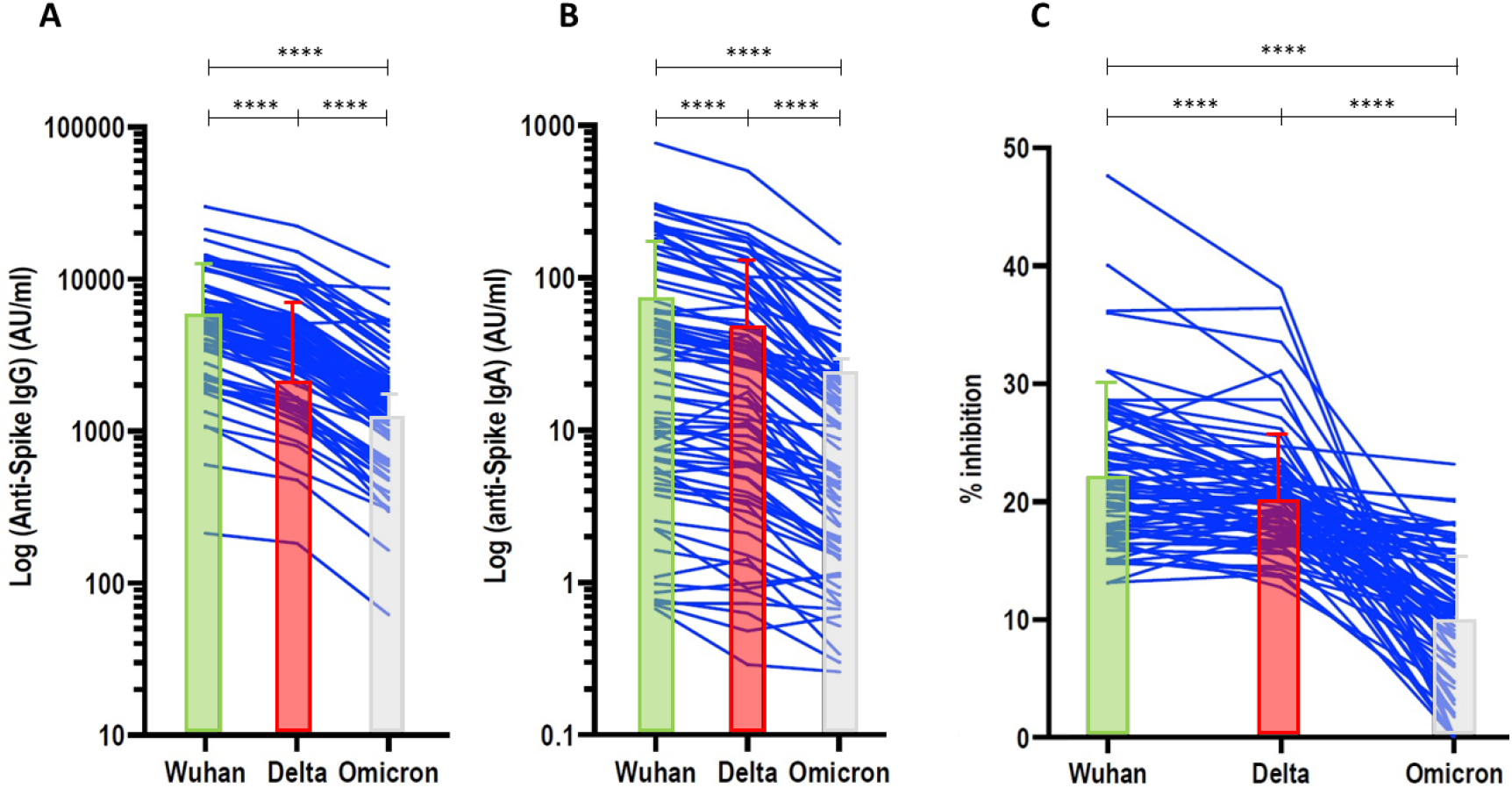
Levels of NELF IgG and IgA to Spike and ability of NELF antibodies to inhibit the binding of Spike to ACE-2. NELF were analyzed for IgG **(A)** and IgA **(B)** to Spike of the ancestral Wuhan strain and its B.1.617.2 (Delta) and B.1.1.529 (Omicron) variant. Data are expressed in arbitrary units per ml (AU/ml) in individual subjects after normalization total IgG and IgA respectively. Medians with the interquartile range are shown. **(C)** NELF were analyzed for inhibiting the binding of Spike of the Wuhan strain and of its variants to ACE-2. Percentages of inhibition in individual subjects are shown. Samples from the same subject are connected by lines. The Wilcoxon matched paired signed rank t test was used to compare the binding inhibition of IgG and IgA to each of Spike of the indicated strain to ACE-2. ****, p < 0.0001.

Our results show that Omicron escapes vaccine-induced mucosal antibody response more efficiently than Delta. This may explain the increased risk of onward transmission of Omicron, consistent with its successful global displacement of Delta in countries with a high vaccination coverage.

## Data Availability

All data produced in the present study are available upon reasonable request to the authors

## Acknowledgements

The authors thank the subjects who volunteered in this study, the medical and paramedical personnel involved in their recruitment and follow-up.

